# Development and validation of a deep learning model for the automated detection of vertebral artery calcification on non-contrast head-and-neck computed tomography

**DOI:** 10.64898/2026.03.15.26348421

**Authors:** Yoko Ueda, Takashi Okazaki, Hiroshi Isome, Aman Patel, Takashi Ichimasa, Rieko Asaumi, Taisuke Kawai, Kaori Suyama, Shogo Hayashi

## Abstract

**Background:** Vertebral artery calcification (VAC), a critical indicator of cerebrovascular disease, is often overlooked in head-and-neck imaging. Manual detection is time-consuming and prone to inter-observer variability. This study aimed to develop and validate a deep learning model for automated detection and quantitative risk assessment of VAC in non-contrast head-and-neck computed tomography (CT) images, bridging the diagnostic gap between dentistry and vascular medicine.

**Methods:** We developed a deep learning model based on the ResNet-18 architecture, designated as Grayscale ResNet, optimized for single-channel CT images. The development followed a two-phase strategy: initial training on 539 axial images from head-and-neck CT image followed by iterative refinement (fine-tuning) using a targeted dataset of clinically significant cases to ensure generalizability. The model’s performance was evaluated using patient-level Receiver Operating Characteristic (ROC) analysis and saliency map visualization for clinical interpretability.

**Results:** The optimized model demonstrated a robust performance in distinguishing between cases with and without VAC. In the independent cohort, the model achieved an area under the curve (AUC) of 0.846. At a specific threshold value (98.6%), the system yielded a sensitivity of 80.0% and a specificity of 90.6%. A saliency map analysis confirmed that the model consistently focused on anatomically relevant vascular regions.

**Conclusions:** The proposed automated system provides an accurate and reliable method for VAC screening using routine head-and-neck CT scans. By transforming incidental imaging findings into a quantifiable risk index, this tool can serve as a vital decision-support system for dentists and radiologists, facilitating early patient referrals and contributing to global stroke prevention.

## Background

Stroke, a leading cause of global mortality and long-term disability, remains a global health challenge [1] The primary cause of ischemic stroke is cervicocranial artery atherosclerosis [2] Vertebral artery calcification (VAC) is an increasingly recognized marker of advanced atherosclerosis [3]. As a key vessel supplying the posterior circulation to the brain, calcification within the vertebral artery can lead to vascular stenosis and occlusion, increasing the risk of vertebrobasilar insufficiency and ischemic stroke [4]. Therefore, early identification of calcified plaques is crucial for timely risk assessment and intervention.

This clinical reality presents a unique opportunity for dentistry. Head-and-neck computed tomography (CT), including cone-beam CT (CBCT), is routinely performed in university hospitals and general clinics for implantology, orthodontics, and surgery. The field of view in these scans often includes the cervical spine. While incidental findings of internal carotid artery (ICA) calcification have been reported [5], the following critical barrier remains: since dentists and oral surgeons are typically not trained to interpret vascular calcification, these significant findings are often overlooked. Furthermore, the diagnostic utility of ICA in CBCT is often limited by artifacts, such as metal prostheses, and by misleading structures, such as the hyoid bone and pharyngeal stones.

In contrast, the vertebral artery, particularly in the upper segments between the occipital bone and the atlas (C1 vertebra), is frequently included in the field of view. This region is generally less obscured by artifacts, presenting a more reliable and clinically relevant target for opportunistic screening than the ICA. Although several studies have examined AI for ICA detection [6–10], data on the use of automated VAC detection on head-and-neck CTs remain lacking.

Therefore, this study aimed to develop and validate a machine-learning model to assist in the automated detection of vertebral artery calcification on head-and-neck CT images. We hypothesized that a tool built on a convolutional neural network (CNN) with the ResNet-18 architecture could serve as a robust adjunct for dentists. The aim is to turn these often-overlooked incidental findings into useful clinical information, enabling quick patient referrals for detailed cardiovascular assessments and directly supporting stroke prevention.

## Methods

### Study Population and Image Acquisition

This retrospective study was approved by the Institutional Review Board of Numazu City Hospital (Approval No. 2023-014). We retrospectively identified 91 adult patients who underwent head-and-neck CT image at the Department of Dentistry and Oral Surgery at Numazu City Hospital between September 2013 and September 2023 (**Table 1**). All examinations were performed using a multi-detector CT system (NM/CT 870 DR, GE HealthCare).The acquisition protocol included a tube voltage of 120 kV and automated tube current modulation (Smart mA). Although the images were initially acquired with a slice thickness of 0.625 mm, they were reformatted into 3 mm slices to create a standardized dataset for AI training. These images were reconstructed using a high-spatial-frequency kernel (Bone Algorithm) to maximize the clarity of calcified lesions and were exported in DICOM format (FOV: 250 mm).

**Table 1.** Distribution of study participants (n = 91) by age group, sex, and the presence of vertebral artery calcification.

| Age Group | Total Participants | Male | Female | Participants with VAC | Male with VAC | Female with VAC |
| --- | --- | --- | --- | --- | --- | --- |
| < 20s | 16 | 7 | 9 | 0 | 0 | 0 |
| 30s | 6 | 2 | 4 | 0 | 0 | 0 |
| 40s | 12 | 2 | 10 | 0 | 0 | 0 |
| 50s | 21 | 10 | 11 | 0 | 0 | 0 |
| 60s | 17 | 12 | 5 | 1 | 1 | 0 |
| 70s | 6 | 3 | 3 | 0 | 0 | 0 |
| 80s | 12 | 6 | 6 | 4 | 2 | 2 |
| 90s | 1 | 0 | 1 | 1 | 0 | 1 |
| **Total** | **91** | **42** | **49** | **6** | **3** | **3** |

Data are shown as the number of individuals in each category. Vertebral artery calcification was observed in six participants (three males and three females), ranging from the 60s to 90s age groups.

The presence or absence of VAC at the C0-C1 level was determined by the consensus of two board-certified oral and maxillofacial radiologists, following previously established criteria [11] This expert consensus served as the ground truth label.

### Phase 1: Initial Dataset and Preprocessing

To standardize the input data and minimize the computational load, a preprocessing pipeline was implemented (**Fig. 1**). All DICOM images were converted into a single-channel grayscale format and resized to 224×224 pixels. Grayscale normalization was applied (mean = 0.5, SD = 0.5) to ensure numerical stability.

**Figure 1.**
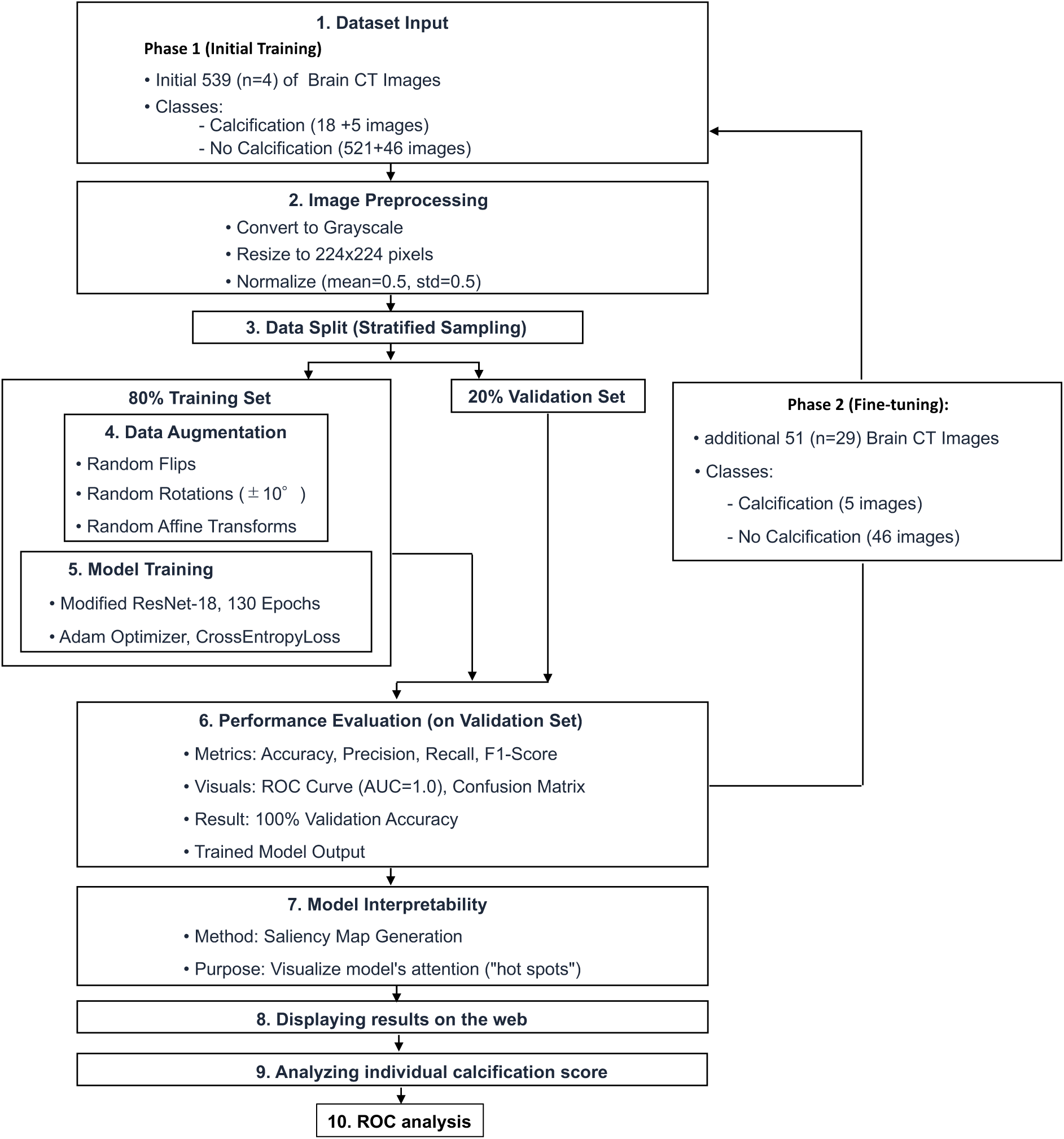
Methodology pipeline for the development and optimization of the AI-based vertebral artery calcification detection system. Illustrated is the complete workflow from initial dataset acquisition to the final ROC analysis (steps 1–10). This process was divided into two strategic phases. Phase 1 (Steps 1–6): Initial training of the prototype modified ResNet-18 model using 539 images from 4 subjects. Although this model achieved 100% validation accuracy and an AUC of 1.0 (Step 6), it exhibited limited generalizability to unseen clinical data. Phase 2 (Step 6 back to Step 1): Iterative refinement through fine-tuning using a targeted dataset of 51 images from 29 patients (Step 6, lateral arrow). These images were selected to include the difficult-to-discriminate clinical features. This two-phase approach facilitates the final calculation of individual calcification scores and a robust ROC analysis (Steps 9 and 10).

For initial model development (Phase 1), we selected four patients (two with VAC and two without) from the cohort. From these scans, 539 axial images containing relevant cervical anatomy were categorized into two classes: calcification (n = 18) and No Calcification (n = 521), because calcification is focal and appears on only a few slices per patient (**Fig. 2A-C**, **Fig. 4A,B**).

**Figure 2.**
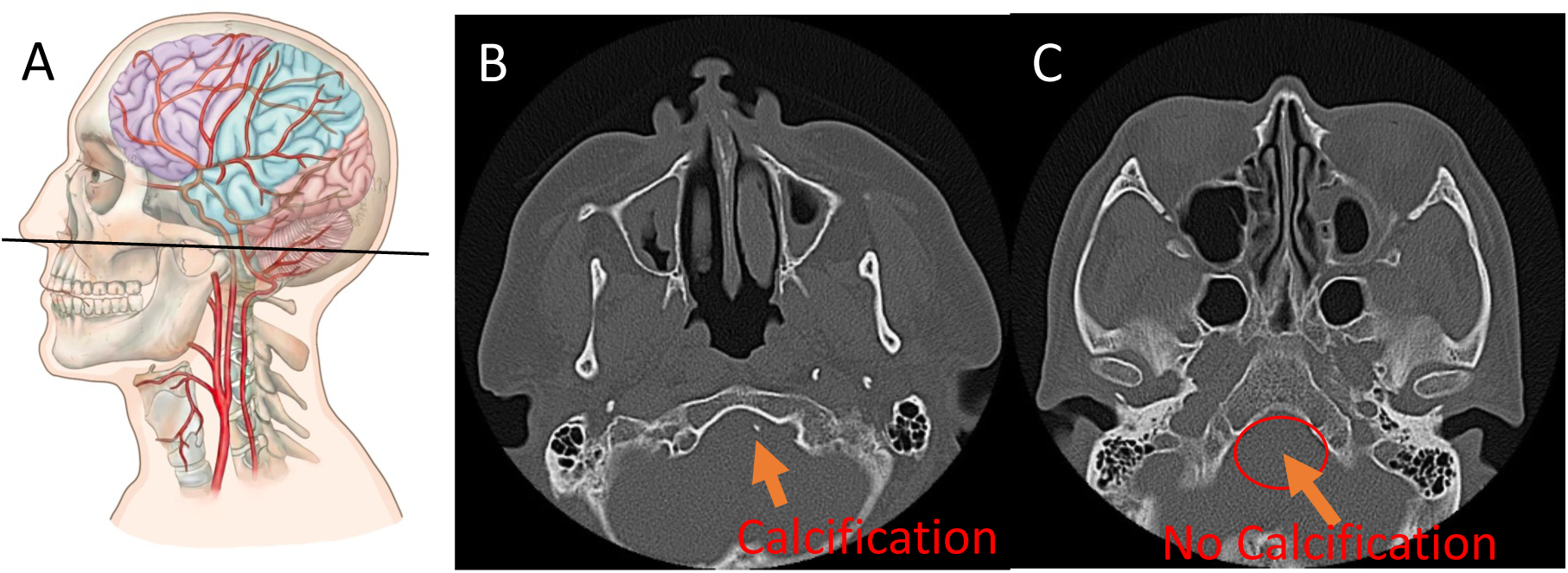
Anatomical landmarks and representative axial CT images at the C0-C1 level. (A) Schematic diagram illustrating the cervicocranial vasculature and the course of the vertebral artery. The black line indicates the axial section plane. (B) Non-contrast axial CT image from a patient with vertebral artery calcification (VAC), identified as a focal hyperdense region (orange arrow). (C) Axial CT image from a patient without VAC. The red ellipse highlights the expected anatomical position of the vertebral artery, demonstrating an absence of calcifiable plaque.

Data augmentation was applied exclusively to the training set to tackle class imbalance (approximately 1:29) and enhance generalization. (**Fig. 1**). Techniques included random horizontal flipping, rotation (±10°), and affine transformations (5% translation). However, the validation set is not augmented.

### Model Architecture and Training

We employed a modified ResNet-18 architecture (GrayscaleResNet) implemented using the PyTorch framework [12,13]. A Residual Neural Network (ResNet) is characterized by skip connections that allow gradients to flow more easily during training, enabling effective learning of deep features without the vanishing gradient problem [12]. We selected the ResNet-18 model from the Torchvision library [13] for its specific advantages in this clinical context. The ResNet-18 model offers a significantly faster inference speed than deeper architectures (e.g., ResNet-50 or 101) while maintaining high accuracy, making it ideal for real-time clinical applications where rapid result delivery is essential.

The model utilizes a pretrained ImageNet backbone with the first convolutional layer adapted for grayscale inputs. The model was trained for 130 epochs using Google CoLab (Tesla T4 GPU). Key hyperparameters included a batch size of 16, a learning rate of 0.001 (with the ReduceLROnPlateau scheduler), and the Adam optimizer. Training performance, including Loss and Accuracy, was monitored to ensure convergence (**Fig. 3A, B**).

**Figure 3.**
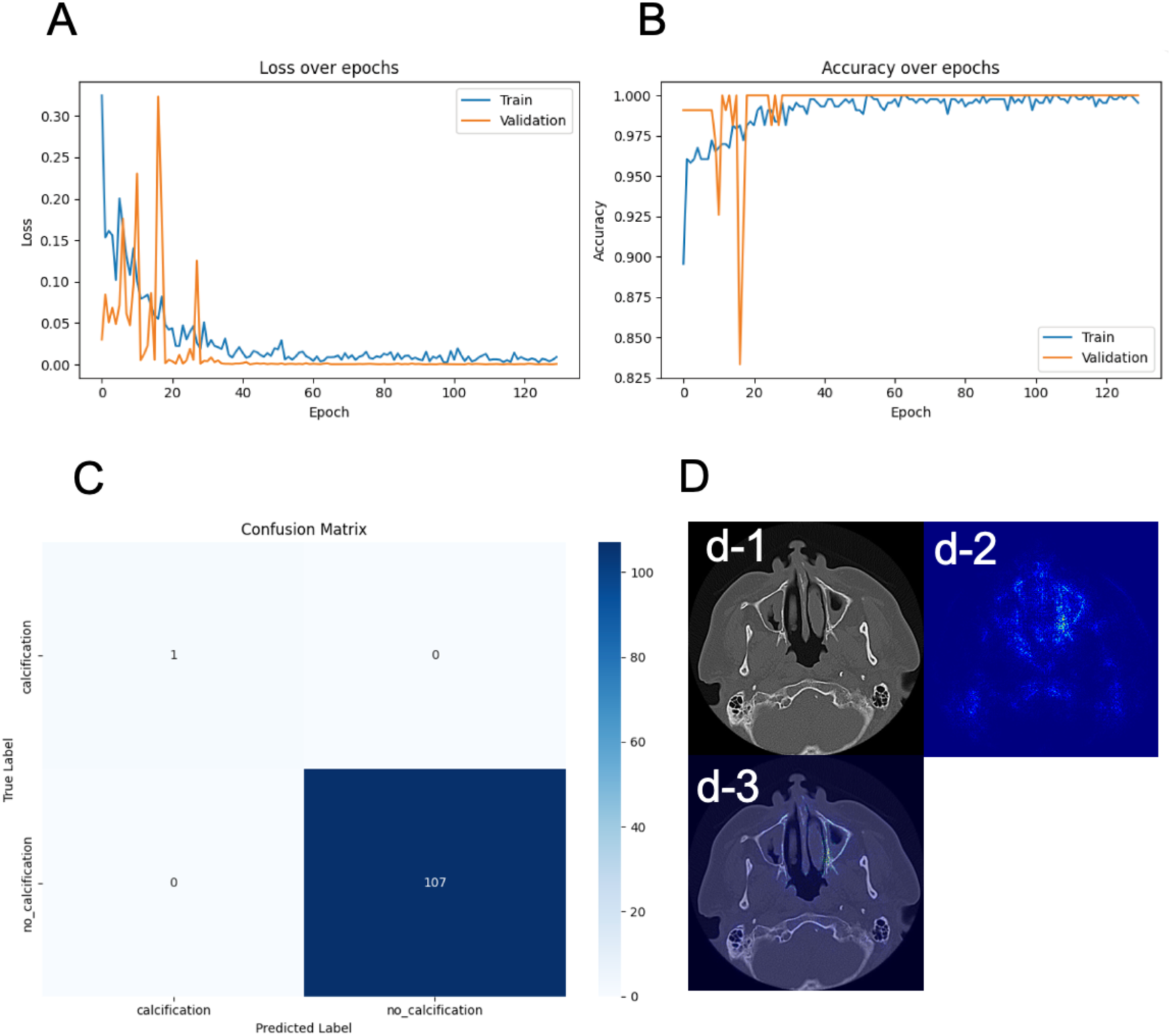
Training performance metrics and model interpretability analysis. (A) Loss over epochs: The graph shows the rapid convergence of both the training (blue) and validation (orange) losses toward zero, with stabilization after approximately 40 epochs. (B) Accuracy over epochs: The model achieved near-perfect validation accuracy (1.00) early in training, indicating robust learning on the initial dataset. (C) Confusion Matrix: Summarizes predictions on the Phase 1 validation set (n = 108). The model correctly classified the positive case and 107 negative cases with no false results. (D) Saliency Map Analysis: Visualizes the model’s decision-making process through gradient-based localization. d-1: Original input image. d-2: Saliency map highlighting influential pixels. d-3: Overlay visualization confirming that the hot spots align with the anatomical location of the vertebral artery, ensuring the model avoids reliance on confounding artifacts.

### Phase 2: Iterative Model Refinement

The initial Phase 1 model showed near-perfect metrics on its internal validation set; however, this was potential overfitting owing to limited patient diversity. Therefore, Phase 2 was initiated to perform fine-tuning using targeted 51 images from a subdataset derived from 29 additional patients selected from the remaining 86 subjects. These 51 images have critically important misleading diagnostic features, such as the odontoid process of the axis and the bony spine. The model was retrained on this expanded dataset to improve discrimination of difficult clinical cases (**Fig. 1**).

### Probability and Risk Score Calculation

To quantify diagnostic certainty for each axial slice, the model used a probability-based scoring system derived from the network’s final output. This process involved three primary steps.

#### 1. SoftMax Normalization

Following the forward pass of the ResNet-18 architecture, the model generated raw, unnormalized scores (logits) for the two target classes: Calcification and No Calcification. These logits are transformed into probabilities using the SoftMax function:

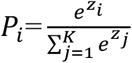

where 𝑍*_i_* represents the logit of class𝑖. This ensures that the probabilities for both classes sum to 1.0 and that each value lies between 0.0 and 1.0.

#### 2. Confidence Extraction

The model’s confidence level was defined as the maximum probability across the two classes.

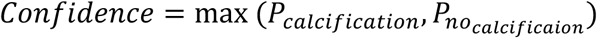

In this binary classification framework, confidence values typically range from 0.5 (maximum uncertainty) to 1.0 (maximum certainty).

#### 3. Quantitative Risk Score (Calcification Index)

The Risk Score (0–100) was calculated from the confidence level to establish a continuous index suitable for the clinical risk assessment and Receiver Operating Characteristic (ROC) analysis. This score represents the likelihood of calcification, as follows:

If Prediction = Calcification:

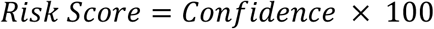

By using this inverse calculation for negative predictions, the system produces a linear scale, where a score near 100 indicates a near-certain presence of calcification, and a score near 0 indicates a near-certain absence.

For the patient-level evaluation, the highest Risk Score across all consecutive axial slices was used as the representative calcification level (**Fig. 4A, B**).

**Figure 4.**
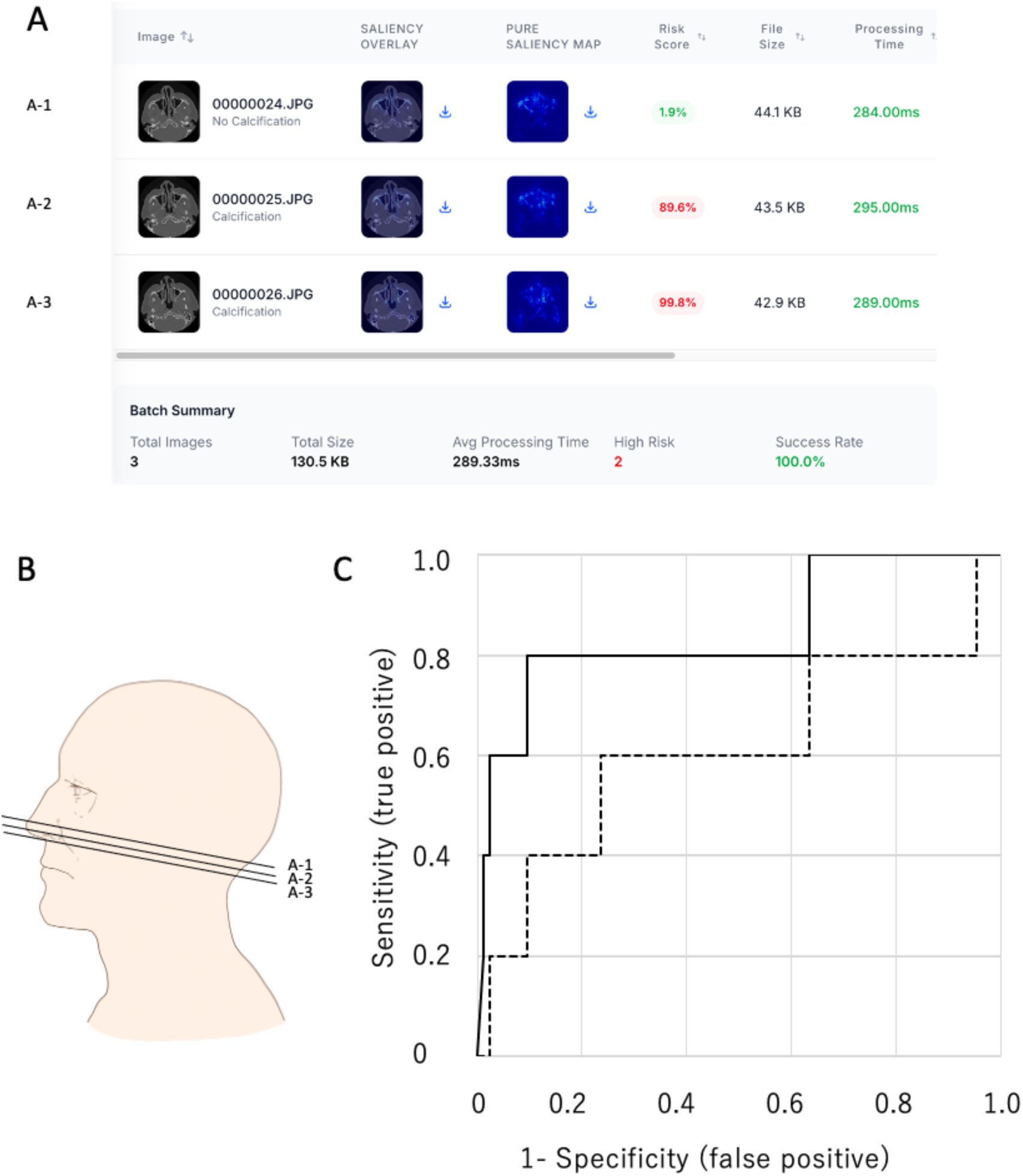
Receiver operating characteristic (ROC) curves for patient-level evaluation and comparing model performance. (A) Automated diagnostic output via the web-based interface. The system calculates a quantitative risk score (Calcification Index) for three typical slices: A-1 (1.9%), A-2 (89.6%), and A-3 (99.8%). For patient-level evaluation, the highest score among all serial sections, e.g., 99.8%, is selected as the representative value for that individual. (B) Schematic illustration of serial axial CT sections of the head and neck for A-1, A-2, and A-3. The AI system evaluates every consecutive slice to ensure comprehensive screening of the vertebral artery. (C) Receiver operating characteristic (ROC) curves comparing model performance. Solid Line (Initial Model): Performance of the Phase 1 prototype (AUC = 0.612). The model exhibited limited generalization when applied to the broader patient cohort. Dashed Line (Final Model): Performance after Phase 2 fine-tuning with 51 targeted clinical images. The optimized model achieved a significantly higher AUC (0.846), with an optimal sensitivity of 80.0% and specificity of 90.6%, confirming its efficacy as a robust screening tool for vertebral artery calcification.

### Web-Based Diagnostic Interface

We developed a web-based prototype to facilitate rapid testing and demonstrate its potential clinical utility. This platform allows users to upload CT images and receive immediate automated diagnostic predictions, with classification results reflecting probability, risk score, and confidence scores displayed directly in the browser (**Fig. 4A**).

### Patient-Level Evaluation and Performance Metrics

The final AI system was used to evaluate the presence or absence of VAC in all 91 participants (**Fig. 1**). For this assessment, all consecutive axial CT slices from each patient’s head and neck scans were processed using this algorithm. The AI calculates the SoftMax probability for each slice. Specifically, slices with a zero probability of calcification were excluded, and the peak value among the remaining positive-probability scores was designated as the patient’s representative calcification level (risk score). This maximum value was subsequently used as the continuous variable to construct the ROC curve and determine the optimal threshold for binary classification at the individual patient level. These representative scores were used to construct Receiver Operating Characteristic (ROC) curves to compare the performance of Phase 1 and Phase 2 (**Fig. 5**).

An optimal threshold was determined based on these scores, and the presence or absence of VAC was subsequently classified as binary (present vs. absent). Standard performance metrics, including accuracy, precision, recall (sensitivity), and F1-score, were calculated based on patient-level results.

### Model Interpretability

To ensure transparency and clinical explainability (explainable AI), we implemented saliency map analysis (**Fig. 4B**). This gradient-based technique highlights pixels that influence the model’s decision, confirming that the model focuses on the vertebral artery rather than on artifacts. Finally, a web-based interface was developed to demonstrate the potential clinical workflow integration (**Fig. 6**).

## Results

### Initial Model Performance (Phase 1)

The performance of the initial prototype (Phase 1) was first evaluated using a held-out internal validation set comprising 20% of the preliminary dataset (108 images). In this subset, the model demonstrated exceptionally high metrics. The detailed performance metrics are listed in Table 2. In this validation set (n = 108), the model achieved a precision, recall, and F1-score of 1.00 for the positive class. The confusion matrix **(Fig. 3C)** confirmed these results, showing no false positives or negatives during the initial phase.

**Table 2.** Precision, recall, F1-score, and support for each class (Calcification, No Calcification)

| Class | Precision | Recall | F1-Score | Support |
| --- | --- | --- | --- | --- |
| Calcification | 1.00 | 1.00 | 1.00 | 1 |
| No Calcification | 1.00 | 1.00 | 1.00 | 107 |

### Iterative Refinement and Final Performance (Phase 2)

Despite the promising results of the internal validation set, subsequent testing of a broader cohort of 87 additional participants revealed significant generalization issues. As illustrated by the solid line in Figure 4C, the initial model yielded an AUC of only 0.612 (cutoff value: 81.4%; sensitivity: 60.0%; specificity: 76.4%) for this expanded dataset, rendering it unsuitable for reliable clinical applications **(Fig.4C)**. The AUC was computed on the held-out test set (the remaining images), not on the training set.

To address these limitations, we implemented a Phase 2 fine-tuning process using a targeted set of 51 critically important diagnostic images. This refinement substantially improved the model’s discrimination capability. The final optimized model, illustrated by the dashed line in Figure 4C, achieved a significantly higher AUC (0.846). At an optimal cutoff of 98.6%, the model demonstrated a sensitivity of 80.0% and a specificity of 90.6%, indicating diagnostic reliability suitable for screening **(Fig. 4C)**.

### Model Interpretability

To verify clinical relevance and ensure explainability, a saliency map analysis was performed on the correct predictions. As shown in Figure 3D, the focal attention of the model (hotspots) consistently aligned with the anatomical location of the vertebral artery **(Fig. 3D)**. This confirms that the classification decisions were based on relevant biological features associated with calcification rather than on those associated with confounding image artifacts.

## Discussion

### Overview and Model Performance

In this study, we developed and evaluated an AI model for automated detection of calcifications in non-contrast head-and-neck CT scans. Our primary goal was to assess the feasibility and performance of this model as a potential diagnostic tool. The results indicate that our approach is highly effective and demonstrates a strong capability to distinguish between images with and without VAC.

The implementation of our iterative refinement strategy proved pivotal in improving the model’s generalizability. The final optimized model achieved an AUC of 0.846 with an optimal probability cutoff of 98.6%. At this threshold, the system demonstrated a sensitivity of 80.0% and specificity of 90.6%. This substantial performance improvement indicates a high level of diagnostic reliability suitable for screening applications, as evidenced by a comparative ROC analysis **(Fig. 4C)**.

The application of artificial intelligence (AI) in medical imaging has significantly evolved, particularly in the detection of calcifications across various modalities. Early efforts to detect carotid artery calcifications (CACs) on panoramic radiographs (PRs) used methods such as fuzzy image contrast enhancement, achieving a detection rate of only 50% [14]. The performance improved with the incorporation of a support vector machine (SVM) to reduce misdetections, which decreased false positives by 75% [15]. Subsequent rule-based approaches combined with SVM achieved a sensitivity of 90%, albeit with 4.3 false positives per image [6]. The last decade has seen a paradigm shift towards deep learning (DL), especially with Convolutional Neural Networks (CNNs), which perform advanced image analysis by learning features directly from image data [16, 17]. For CAC detection in PRs, models such as Faster R-CNN reported an accuracy of 83% [18], whereas more advanced cascaded networks achieved an AUC of 0.996 and an F1-score of 88.8% [8,9].

Similar advancements have been reported for cone-beam computed tomography (CBCT). A pioneering study applied a deep learning pipeline using Inception V3 and U-Net, achieving a diagnostic accuracy of 96.35% for CAC detection [19]. More recent U-Net-based models have segmented extracranial and intracranial carotid atheromas with high sensitivities of 92% and 96%, respectively [9]. In CT imaging, deep learning has also been used to detect intracranial carotid artery calcification with high sensitivity (83.8%) and positive predictive value (88%) [20].

Despite these successes in carotid imaging and the growing histological evidence regarding the prevalence of medial calcification in vertebral arteries [21,11], effective AI models specifically optimized for Vertebral Artery Calcification (VAC) on CT remain rare. Notably, Mahdian et al. [10] reported lower detection performance of VAC was than extracranial (ECC) and intracranial carotid calcification (ICC). The authors attributed this performance gap primarily to the limited training data available for VAC (only 21 cases, compared with 74 for ECC and 40 for ICC). Furthermore, they noted that the minute size and subtle features of VACs made it difficult for algorithms to accurately segment them from surrounding tissues, leading to lower segmentation performance metrics for segmentation-based approaches.

Compared with the existing literature, our model’s performance is highly competitive. The primary strengths of our study were the simplicity and effectiveness of the ResNet-18 architecture. Unlike complex, multilayered pipelines that rely on the precise segmentation of regions of interest (ROI), a task known to fail with small, subtle targets, our approach uses direct classification. As illustrated in Figure 1, we relied on careful preprocessing rather than fragile segmentation engines **(Fig.1)**. This design choice bypasses the difficulties noted by Mahdian et al., enabling future functional expansion and ensuring model generalizability.

Second, our decision to focus specifically on the anatomical region between the occipital bone and the atlas (C1 vertebra) likely contributed to the model’s high performance. The vertebral artery (VA) in this region represents a critical junction and transition point, making it susceptible to mechanical compression/injury and intrinsic vascular disease [22]. This area encompasses the V3 segment (suboccipital) and the entry point of the V4 segment (intracranial) [23]. Calcifications in the vertebral artery often manifest radiographically at the level of the foramen magnum as circular or semicircular masses between the V3 and V4 segments [11] (Fig.2B). By targeting this specific high-risk area rather than attempting to analyze the entire vertebral artery, which originates from the subclavian artery and traverses a complex path into the skull, we achieved a high detection rate with reduced background noise.

Despite these promising results, this study had several limitations. First, this was a retrospective study using data from a single institution, and variations in imaging conditions arose from using different devices. Therefore, the model’s performance on external datasets from other scanners or diverse patient populations was not determined. Second, the ground truth for this study was based on visual assessment by an imaging specialist, rather than on a more detailed examination, such as contrast-enhanced CT. Finally, our current model primarily performs binary classification. However, as detailed in the Materials and Methods, we have already established a mathematical foundation for a Quantitative Risk Score (Calcification Index).

In future studies, we intend to validate this model in a multicenter, prospective cohort study. We aimed to develop this tool from a simple binary classifier into a quantitative system capable of calculating a ‘calcification score’ or index, similar to the Agatston score used in cardiac imaging. This would allow clinicians to assess the presence and the severity of risk. To mitigate confounding from other calcified pathologies that may be incidentally detected (e.g., pharyngeal stones, thyroid calcifications, or salivary stones), future iterations of the model could be expanded to address a multiclass classification problem. This would help to differentiate between calcification types and provide more accurate diagnostic information. Furthermore, by exploring model interpretability with techniques such as Grad-CAM and incorporating clinical data (e.g., age, sex, and medical history) into machine learning models such as Random Forest, we can better explain the model’s decision-making process and increase clinical trust. Additionally, we intend to integrate AI-generated outputs with electronic health records to provide a more holistic patient assessment.

## Conclusions

The AI model presented in this study demonstrates outstanding accuracy, precision, and recall for the automated detection of calcification in vertebral arteries. Applying AI with high accuracy may enable early risk assessment of cerebrovascular disorders. The findings suggest that this tool has significant potential for integration into the clinical workflow as a decision support system for radiologists. Flagging suspicious cases can help to streamline the diagnostic process, reduce oversight errors, and ultimately contribute to earlier and more accurate disease detection. Although further validation and refinement are necessary prior to clinical implementation, this study makes a significant contribution toward achieving this goal. By facilitating the early detection of VAC in dental settings, this tool has the potential to significantly reduce the burden of stroke and cardiovascular disease at the population level.

## List of Abbreviations

VAC: Vertebral artery calcification
CT: computed tomography
ROC: Receiver Operating Characteristic
CNN: convolutional neural network
PRs: panoramic radiographs
AI: artificial intelligence
CACs: carotid artery calcifications
CBCT: cone-beam computed tomography
CAC: Carotid Artery Calcification
ICC: Intracranial Carotid Calcification
ECC: extracranial Carotid Calcification
EHR: electronic health records

## Declarations

### Ethics approval and consent to participate

This study was approved by the Institutional Review Board of Numazu City Hospital (Approval No. 2023-014). Due to the retrospective nature of the study, which used anonymized imaging data, the requirement for informed consent was waived by the Institutional Review Board of Numazu City Hospital. All methods were performed in accordance with relevant guidelines and regulations.

### Consent for publication

Not applicable.

### Availability of data and materials

The datasets generated and/or analyzed during the current study are not publicly available because of patient privacy restrictions but are available from the corresponding author upon reasonable request.

### Authors’ contributions

YU and SH conceived and designed the study. YU, RA, and TK collected and curated imaging data. HI, AP, and TI developed the machine-learning model and performed the data analysis. TO provided expert radiological evaluations and established ground-truth labels. KS and SH supervised the study. HI and YU drafted the manuscript. All the authors have read and approved the final version of the manuscript.

## Acknowledgements

We thank the radiologists at Numazu City Hospital for their invaluable assistance in establishing the ground truth of the image dataset.

